# Scaling behaviors of deep learning and linear algorithms for the prediction of stroke severity

**DOI:** 10.1101/2022.12.05.22283102

**Authors:** Anthony Bourached, Anna K. Bonkhoff, Markus D. Schirmer, Robert W. Regenhardt, Martin Bretzner, Sungmin Hong, Adrian V. Dalca, Anne-Katrin Giese, Stefan Winzeck, Christina Jern, Arne G. Lindgren, Jane Maguire, Ona Wu, John Rhee, Eyal Y. Kimchi, Natalia S. Rost, the MRI-GENIE and GISCOME Investigators and the International Stroke Genetics Consortium

## Abstract

**Introduction:** Deep learning has allowed for remarkable progress in many medical scenarios. Since deep learning prediction models often require 10^5^-10^7^ examples, it is currently unknown whether deep learning can also enhance predictions of symptoms post-stroke in real-world samples of stroke patients that are often several magnitudes smaller. Such stroke outcome predictions however could be particularly instrumental in guiding acute clinical and rehabilitation care decisions. We here compared the capacities of classically used linear and novel deep learning algorithms in their prediction of stroke severity.

**Methods:** Our analyses relied on a total of 1,430 patients assembled from the MRI-GENIE collaboration and a Massachusetts General Hospital-based study. The outcome of interest was NIHSS-based stroke severity in the acute phase after ischemic stroke onset, which we predict by means of MRI-derived lesion location. We automatically derived lesion segmentations from diffusion-weighted clinical MRI scans, performed spatial normalization and included a principal component analysis (PCA) step, retaining 95% of the variance of the original data. We then repeatedly separated a train, validation, and test set to investigate the effects of sample size, we subsampled the train set to 100, 300, and 900 and trained the algorithms to predict the NIHSS score for each sample size with regularized linear regression and an 8-layered neural network. We selected hyperparameters on the validation set. We evaluated model performance based on the explained variance (R-squared) in the test set.

**Results:** While linear regression performed significantly better for a sample size of 100 patients, deep learning started to significantly outperform linear regression when trained on 900 patients. Average prediction performance improved by ∼20% when increasing the sample size 9x (maximum for 100 patients: 0.279 ± 0.005 (R^2^, 95% confidence interval), 900 patients: 0.337 ± 0.006).

**Conclusions:** For sample sizes of 900 patients, deep learning showed a higher prediction performance than typically employed linear methods. These findings suggest the existence of non-linear relationships between lesion location and stroke severity that can be utilized for an improved prediction performance for larger sample sizes.

## Introduction

Recent estimates suggest that ∼12 million people experienced a new stroke worldwide in 2019, while a total of ∼100 million people lived after having experienced a previous stroke that year.^1^ Additionally, stroke is the most burdensome neurological disorder, as highlighted by evaluations of years of full health lost to disability and death.^2,3^

Thus, stroke is both a commonly occurring, as well as a socioeconomically relevant disease and renders any efforts to optimize stroke care exceptionally important. Precision medicine has been a key focus of these efforts in recent years, as it holds promise to optimize patient outcomes. From a methodological standpoint, the realization of this individualized care is particularly linked to the fruitful combination of artificial intelligence and big data.^4^ In fact, many stroke outcome studies have employed classic machine learning algorithms to predict stroke outcomes from various sources of neuroimaging data.^5,6,7^ As such, there has thus been a quick adaptation of novel and powerful methods as soon as computational resources allowed to use them.

The capacity of deep learning for pattern recognition and classification has been especially emphasized for complex and unstructured problems such as chemistry,^8^ physics,^9^ art history,^10–14^ and even human behavior.^15,16^ Similarly, the promises of deep learning for medicine are innumerable. Indeed, some specific biomedical fields have seen major advancements. Examples can be seen in algorithms capable of the prediction of protein structures based on their amino acid sequence (AlphaFold),^17^ histopathological evaluations of tumor tissue^18^ or enhanced automatic medical image processing, relating to preprocessing^19^ and automatic segmentation of pathological brain changes.^20,21,22^

However, the published literature on deep learning-based stroke outcome prediction is comparatively sparse. This may be due to previously relatively small available data set sizes, often only a few hundred subjects, in stroke outcome studies. Two recent studies^23,24^ showed a benefit of deep learning algorithms for the prediction of favorable functional outcome post-stroke. More specifically, both teams trained convolutional neural networks (CNNs) on acute imaging data, that is non-contrast CT data^23^ and MRI-based diffusion-weighted imaging (DWI) data,^24^ and compared the resulting performance with established, basic clinical scores, such as the ASPECT score.^25^ Both studies relied on ∼200-300 patients in total for model derivation and validation. In contrast, Chauhan and colleagues performed a direct comparison of (non-linear) deep learning algorithms and linear algorithms and their capacities to predict language impairments post-stroke based on DWI-derived lesion location information.^26^ They did not find any evidence for a superiority of deep learning, but noted that a combination of deep learning for the refinement of DWI information and ridge regression was most optimal in their specific set-up. Importantly, their analyses were based on a maximum sample size of 132 patients.

There have been substantial dataset size increases in stroke *neuroimaging* studies with available stroke lesion data in recent years. These occurred primarily within the framework of large, international collaborations, such as the Meta VCI map consortium (∼3,000 patients),^27^ ENIGMA (∼2,000 patients),^28^ or MRI-GENIE (∼2,800 patients),^29^ but also in some single center, or national settings, such as University College London Hospital (∼1,300 patients),^5,30^ Hallym University Sacred Heart Hospital, or Seoul National University Bundang Hospital (∼1,400 patients).^31,32^ In addition, stroke is such a common disease that it may well be feasible to acquire even larger datasets. This aspect is exemplified by ongoing studies, such as the DISCOVERY study with a planned inclusion of 8,000 stroke patients.^33^ These increase in dataset sizes allows for new opportunities to test their relevance for the performance of deep learning for stroke outcome predictions.

Prediction performance will conceivably increase with dataset size independent of the algorithm used.^34^ This means that even linear algorithms are expected to improve up to some asymptotic value. In contrast, deep networks are expected to have a higher asymptotic performance value – since the learnable function space for a deep net is a superset of that of a linear model – at the cost of more challenging optimization. These projections may eventually represent further justifications to invest in costly and time-consuming large-scale study endeavors and motivate deep learning-based approaches.

The present study investigates the systematic evaluation of deep learning for the prediction of stroke severity based on neuroimaging-derived lesion location information in a large, multicenter cohort.^29^ While there are categorical differences in how linear and deep learning algorithms are trained and optimized – with deep learning being more complex and generally more difficult to optimize^35^ – we aimed to develop a methodological setup that represented a fair playground for both approaches. To get further insights into the role of sample size, we randomly repeatedly subsampled to three increasing dataset sizes: 100 patients, 300 patients and 900 patients. By these means, we aimed to answer the questions: Are there non-linear effects between the lesion location and stroke severity that could be leveraged by deep learning models? Do larger stroke datasets comprising ∼1,000 patients already represent an advantage over currently primarily available ones in the range of a few hundred patients?

## Methods

### Patient samples

To increase our sample size, we merged data of patients with acute ischemic stroke originating from the multicenter MRI–Genetics Interface Exploration (MRI-GENIE) study,^29^ and a retrospective Massachusetts General Hospital (MGH)-based data set. We included patients with available quality-controlled DWI-based lesion segmentations and information on acute stroke severity, as measured by the National Institutes of Health Stroke Scale (NIHSS, 0-42, 0: no measured deficits, 42: maximum stroke severity). All patients or their proxies of the MRI-GENIE study gave written informed consent in accordance with the Declaration of Helsinki. Given the retrospective character of the MGH-based study, it was performed under a waiver of consent. The study protocols were approved by MGH’s Institutional Review Board (Protocol #: 2001P001186, 2003P000836, and 2013P001024) and the Review Boards of individual sites.

### Neuroimaging data and preprocessing

We utilized each patient’s acute MRI-based diffusion-weighted imaging (DWI) scan (c.f., **supplementary materials** for a description of imaging parameters). DWI-based stroke lesion segmentations were generated by automated routines that differed for the two datasets. In case of MRI-GENIE, these segmentations were produced by means of a validated ensemble of 3-dimensional convolutional neural networks.^36^ In case of the MGH-based study, we employed an in-house deep learning-based algorithm (c.f., **supplementary materials** for details). DWI scans and corresponding lesion segmentations were non-linearly normalized to the common Montreal Neurological Institute (MNI)-space.^37^ To ensure a high quality of both lesion segmentations and spatial transformation, we performed a comprehensive quality control that comprised the manual evaluation of spatially normalized DWI scans in combination with the respective lesion segmentation (three experienced raters: AKB, MB (MRI-GENIE) and JR (MGH-based study)).

Each lesion segmentation comprised binary information for altogether 902,629 voxels. We, therefore, initially performed a dimensionality reduction step, as commonly done in imaging-based stroke outcome studies.^38,39^ We employed principal component analysis (PCA) of the voxel-wise lesion segmentation information and retained as many components as were necessary for explaining 95% of the variance in the lesion data.

### Computational framework and employed algorithms for the prediction of stroke severity

We repetitively separated the entire dataset (total n=1,430) into train, validation and test sets of the sizes 915, 229 and 286, respectively.^4^ The test set comprised approximately 20% of the entire dataset, and the validation set approximately 20% of the remainder, as is a common convention for small datasets.^40^ We repeated this random split into train, validation and test sets 500 times. In case of the train set, we further subsampled to samples of 100, 300 and 900 patients by drawing from the entire sample of 915 without replacement. We inserted this subsampling step with the idea of getting insights on the effect of sample size, as deep learning algorithms are known to result in better performance for larger dataset sizes. We normalized the NIH stroke severity score to be in the range of 0 and 1 by dividing all scores by the maximum of 42. We inserted this preprocessing step to model the outcome as a Bernoulli distribution and utilized a Binary Cross-Entropy (BCE) loss function. We trained algorithms to predict stroke severity via gradient descent using backpropagation relying on Adam optimization.^41^ We opted for a batch size of 64 to enable a fair comparison between training dataset sizes and ran as many batches as were necessary to iterate through all examples for an epoch (therefore, two for datasets of 100 patients and 14 if 900 patients). To optimize model hyperparameters, we repeated the described training procedure for 49 individual models with different combinations of hyperparameter constellations (learning rate = [1.0, 0.3, 0.1, 0.03, 0.01, 0.003, 0.001], weight decay (regularization): [0.1, 0.03, 0.01, 0.003, 0.001, 0.0003, 0.0001]).

We ran the entire analysis pipeline for two different models that varied in the depth of their architecture. As a baseline model that is also the closest to the ones classically employed in stroke outcome prediction studies,^39^ we implemented a *l*_*2*_-regularized logistic linear regression model with a sigmoid activation function. This model therefore corresponded to a 1-layered neural network. The deepest model that we implemented was an 8-layered neural network with seven hidden layers (dimensions: 512, 256, 128, 64, 32, 16, 8, c.f., **Figure 1** for an intuition).

**Figure 1.**
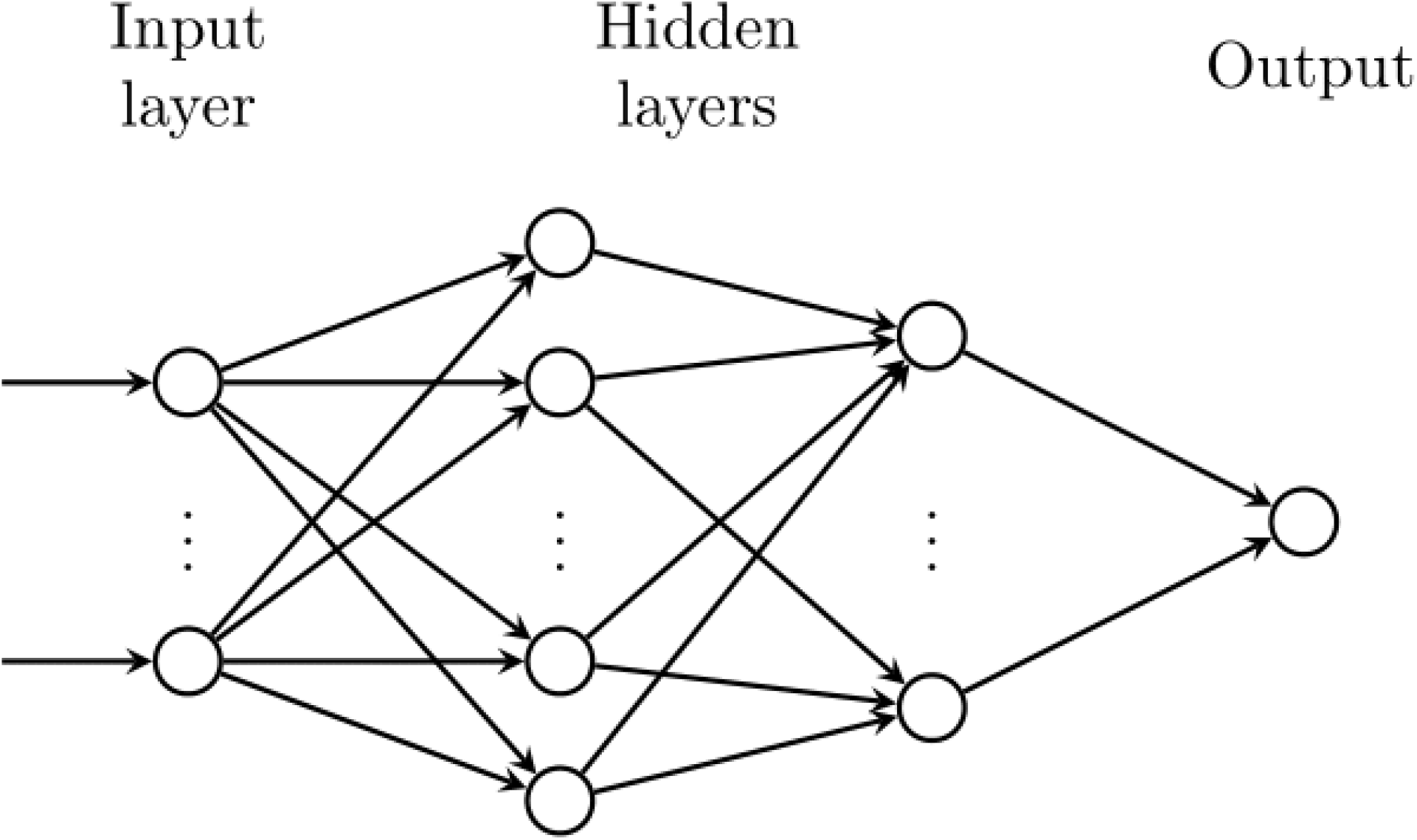
Graphical scheme of the deep learning model. Each hidden layer has ReLU^42^ activation functions, and the output has a sigmoid activation function that squashes the output between 0 and 1.

### Model selection and performance evaluation

We evaluated prediction performance as explained variance based on the coefficient of determination, R-squared. We computed the R-squared value in the test set specifically for the model with the overall highest R-squared value in the validation set. To determine this highest R-squared value in the validation set, we selected the checkpoint in the training process that corresponded to the highest R-squared value in the validation set. That is, we trained for a fixed number and then chose the point with the highest R-squared value, rather than stopping training, when the error stopped decreasing. We report the test set R-squared value as averaged across the 500 random splits of the entire dataset into train, validation, and test sets. Finally, we compared the performance of the two different algorithms with respect to the mean explained variance and linked 95%-confidence intervals. We determined significant differences in prediction performance based on non-overlapping confidence intervals.^30,38^

### Data and code availability

The authors agree to make the data available to any researcher for the express purposes of reproducing the here presented results and with the explicit permission for data sharing by individual sites’ institutional review boards. Prediction analyses were implemented in Python 3.7 (predominantly relying on packages: Pytorch 1.9^43^). Jupyter notebooks will be made available GitHub upon acceptance.

## Results

Our analyses were based on 1,430 patients with acute ischemic stroke (792 MRI-GENIE patients, 638 patients from the MGH-based study). The average age was 66.3 (standard deviation (SD): 15.0) years, and 43.1% were female patients. Patients had a median acute stroke severity of 4 (interquartile range (IQR): 6). The median lesion size was 5.0 ml (IQR: 26.7ml, **Table 1**). **Figure 2** presents a lesion overlap visualization. The highest lesion overlap was located subcortically in middle cerebral artery territory, as well as insular cortex. The PCA-dimensionality reduction step resulted in 504 retained components that served as input to our two algorithms.

**Table 1.**
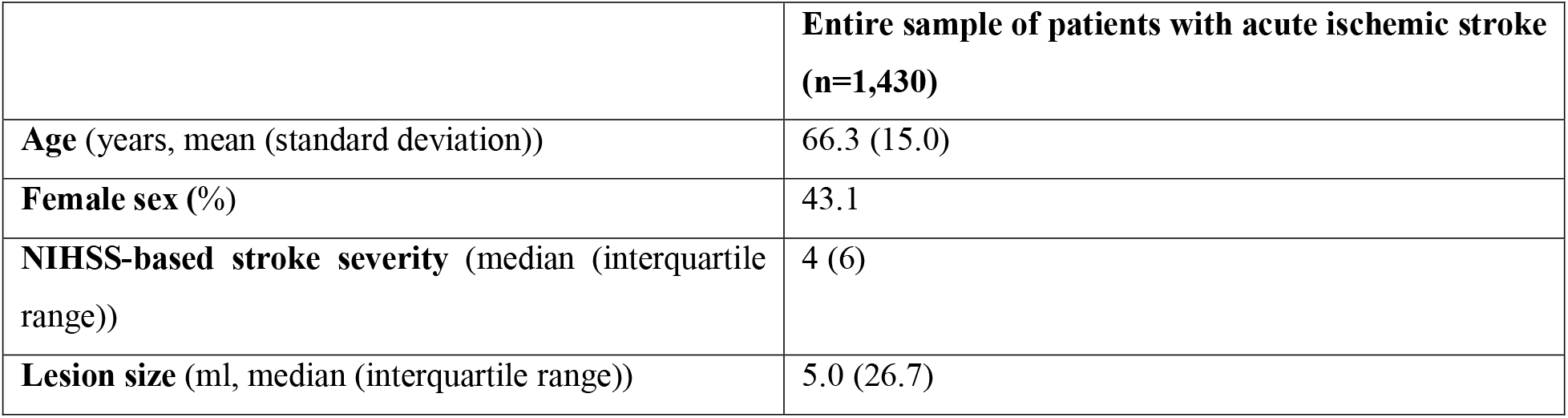
Summary of patient characteristics.

**Figure 2.**
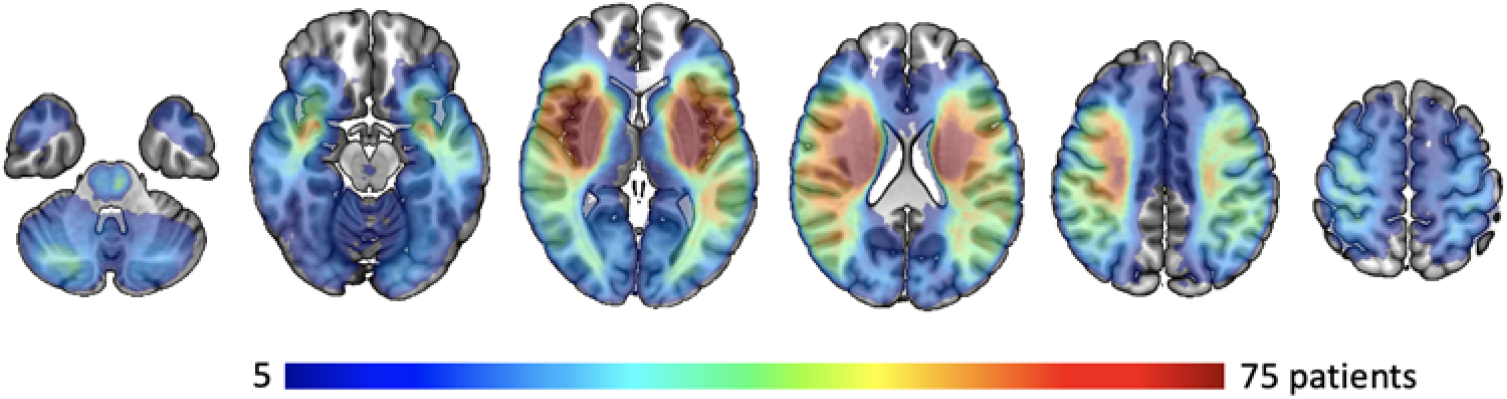
Lesion overlay of all 1,430 patients with acute ischemic stroke. The highest lesion overlap was found in middle cerebral artery territory, more specifically subcortically in proximity to the lateral ventricles and insular cortices of both the left and right hemisphere, which was also expected from prior work.^44,32,45^ Fewer lesions affected bilateral territories of the posterior cerebral arteries (PCA). Our sample furthermore does not provide sufficient coverage of bilateral anterior cerebral artery territories.

### Prediction of stroke severity

#### 100 – 300 – 900 subjects

Linear regression resulted in a significantly higher prediction performance compared to deeper models, when training on the stroke data of 100 patients. The mean performances of the linear regression models totaled 0.279 ± 0.005 (R^2^, 95% confidence interval), compared to 0.250 ± 0.007 for the deep learning models. The non-overlapping 95% confidence intervals thus showed a significant advantage of the linear method for our smallest tested sample size of 100 patients. In case of a training dataset of 300 patients, linear models and deep learning performed similarly, as indicated by their overlapping 95% confidence intervals: The exact mean performances were 0.292 ± 0.006 for linear regression, and 0.296 ± 0.006 for deep learning. The situation of initial superiority was reversed for 900 patients, where deep learning achieved significantly higher prediction performance with an explained variance of 0.337 ± 0.006, compared to the linear model (R^2^ = 0.316 ± 0.006, **Figure 3**).

**Figure 3.**
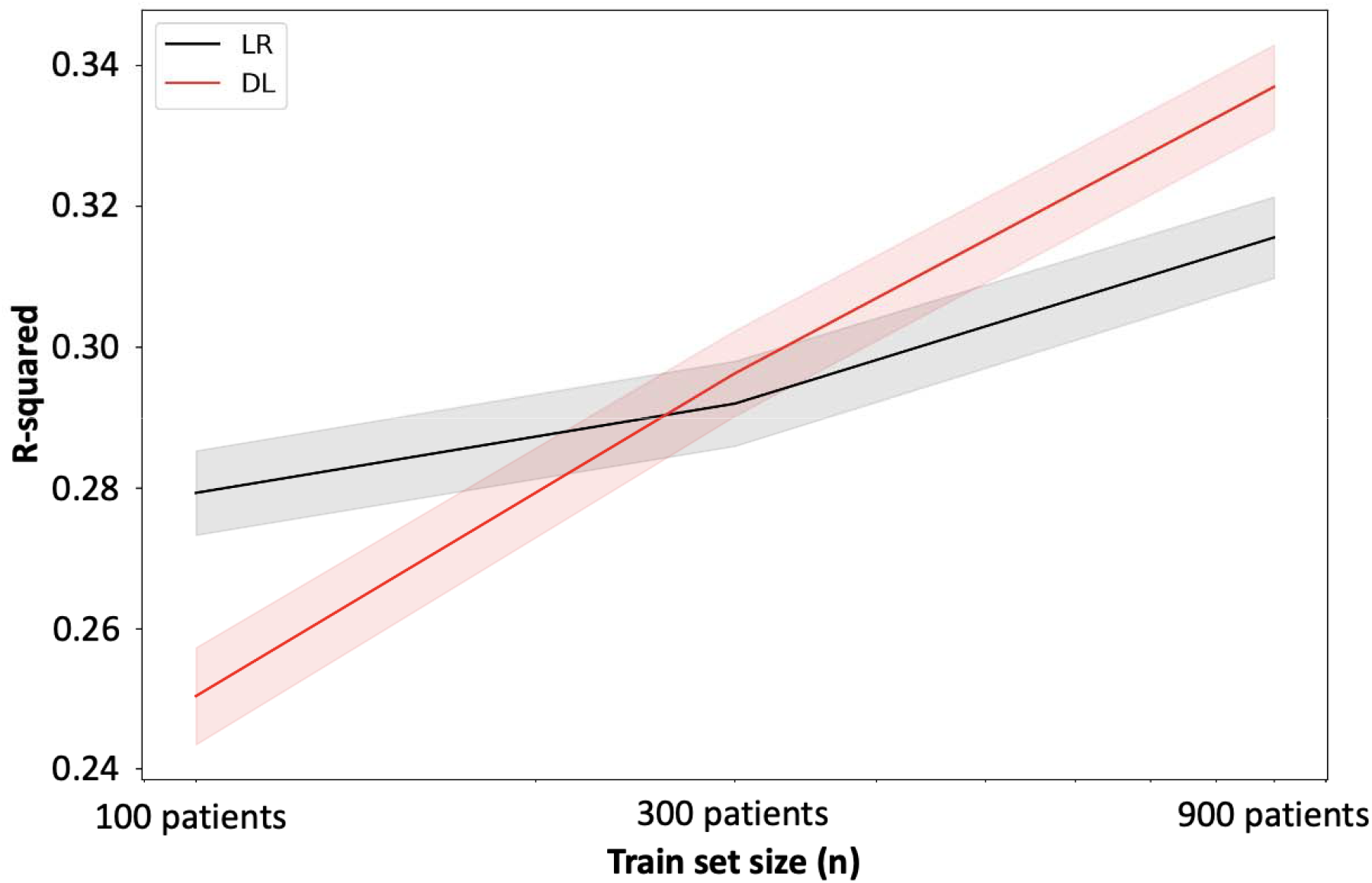
Average prediction performance of stroke severity on the test set over 500 different random data splits in terms of explained variance (R^2^, y-axis) depending on train set size (x-axis). Models were trained for three separate train sample sizes of 100 patients, 300 patients and 900 patients. The x-axis here displays those sizes logarithmically. While the linear regression model performed favorably for a sample size of 100 patients, deep learning started to significantly outperform linear regression when trained on 900 patients. Average predictio performance improved by ∼20% when increasing the sample size 9x.

Altogether, we thus observed that the maximum performance in mean explained variance when going from 100 to 900 patients increased independent of the employed model: by 0.04 for the linear model and by 0.09 for the deep learning model (**Figure 3**).

## Discussion

In this study, we examined the capacity of deep learning for the prediction of stroke severity in relation to linear models and the dependence of training set sample size. Deep learning outperformed more common linear methods for training set sizes of 900 patients with ischemic stroke. Such an advantage was not detectable for sample sizes of 300 patients, which may be seen as an infliction point, as, in fact, the advantage was reversed for sizes of 100 patients. In this case of small training samples, linear methods performed significantly better than deep learning approaches. Independent from this switchover in best performance, there were notable increases in the prediction performance for both linear, as well as deep methods with larger training set sizes.

Our results match the common notion that deep learning-based prediction performs better for larger sample sizes. However, “large” in the context of deep learning-based prediction studies typically refers to samples with 10^5^ to 10^7^ examples, and not 10^2^-10^3^, as in our study. Thus, it may be a particularly encouraging finding that the benefit of deep learning was already appreciable for our only moderately large sample size. Essentially, the significantly higher performance of deep learning compared to linear models suggests that there are non-linear effects between where in the brain a lesion occurs and the severity of stroke symptoms – effects that may only be captured with more flexible models in combination with a sufficiently high number of observations from which to learn. This existence of non-linear effects may be even less surprising when considering that our outcome variable, the NIHSS score, is a global score and combines impairments in several functional systems at once. Given our findings and the fact that there are increasingly more emerging large-scale stroke imaging datasets comprising data of >10^3^ patients with ischemic stroke,^27-32^ it may be promising to include more flexible algorithms, such as deep learning, in the collection of routinely employed algorithms to achieve best possible results for outcome prediction. Similarly, our findings indicate that linear models are best used for small datasets, supporting current practice. Furthermore, linear models provide the highest level of transparency^46^ and may hence be the preferred choice of model at any data set size if the goal was interpretability, rather than prediction.

In addition, the evaluation of maximum performance across smaller to larger sample sizes, especially also the gain from 300 to 900 patients, suggests that it is reasonable to expect further gains with yet larger samples than the currently investigated one. In view of their larger increase in performance from smaller to larger samples, this may be especially true for deep learning models. These projections hence support and justify the ambitions of large, international collaborations aiming to recruit several thousand patients with stroke. It remains to be seen however, whether prediction performances can be increased to an extent that renders them immediately clinically useful. For example, would an increase in prediction performance to an explained variance of 50% be more helpful in clinics than our presented explained variance of 34%, or would clinical utility be given for values of >90% only?

Altogether, several aspects beyond a larger sample size in combination with machine learning techniques and increased prediction performances may influence the clinical utility of future prediction models positively: First, this utility may be directly linked to the importance of the measured outcome scores. Is it an outcome of concrete relevance for patients and their everyday life, such as measure of motor or cognitive impairment? For instance, the Determinants of Incident Stroke Cognitive Outcomes and Vascular Effects on RecoverY (DISCOVERY) study is projected to acquire detailed cognitive scores in 8,000 patients with stroke.^33^ Information on the predicted cognitive level of functioning, as derived from this large sample, may be directly helpful to future patients, their families, and health care professionals, as it will allow for realistic expectations, and optimized, tailored care.^47^

Another relevant consideration is the integration of information from additional sources beyond imaging^48^ – we here solely considered information on the ischemic lesion as apparent on the DWI scan, that represents, essentially, the location, as well as extent of the lesion. Conceivably, the maximum prediction performance will increase with the addition of sociodemographic characteristics and the acute clinical presentation, as well as further imaging-derived information, such as white matter changes,^49,50^ subtle imaging characteristics as captured by radiomics,^51,52^ and compound measures, such as the estimated brain reserve.^53^ Once again, it may be particularly important to explore the potential of combining information from several sources in the context of various sample sizes and algorithms. Most likely, the true benefit may only become apparent for larger samples sizes in combination with models that can capture non-linear effects.

### Strengths and limitations

Our study has several strengths. In addition to the availability of a large data set of ischemic stroke patients with acute imaging available, we exclusively present the prediction performance for a test set. This test set was neither implicated in the training of algorithms, that is the estimation of model weights or optimization of hyperparameters, nor their validation. We therefore adhered to established deep learning standards. Additionally, we paid great attention to create a fair playground for the systematic comparison of deep learning and linear models that are typically both linked to varying optimization and training strategies. We here wrote custom code to optimize our linear models in the same train-validation-test split scenario, as optimal for deep learning. Altogether, we ensured that neither the linear, nor the deep learning model were disadvantaged.

One important limitation of our study is our focus on lesion segmentation information from clinical DWI scans. Given the acquisition in clinical routine, the resolution of these scans was comparably low. Moreover, we employed an initial (linear) principal component analysis (PCA)-step for dimensionality reduction from the image with almost 1 million voxels to 504 principal components.^6,39^ In the future, it may be beneficial to test various linear – and non-linear – approaches for initial dimensionality reduction, as their relevance for prediction performance is currently underexplored. While dimensionality reduction has been an important first step for subsequent training of linear regression algorithms and is commonly performed in stroke outcome studies, deep learning algorithms may be capable of favorably handling voxel-wise data without an intermediate dimensionality reduction step. Another limitation is our focus on a global score, such as NIHSS-based stroke severity. NIHSS subscores were not available to us. However, while the NIHSS is a broad compound score, it might be a valid first step to test the capacity of deep learning in the larger sample size regimen. Especially in view of initiatives that introduce recommendations for enhanced data harmonization between different stroke studies,^54^ future studies testing the validity of our conclusions for specific outcomes, such as motor impairments and cognitive functions, may be feasible.

### Conclusion

We here present first evidence that deep learning can predict stroke severity from lesion information significantly better than linear models once the training set size is sufficiently large (900 patients). Conversely, linear models performed significantly better in case of smaller training samples of 100 patients. Prediction performance generally increased with increasing sample size. In summary, our findings suggest the existence of non-linear relationships between lesion location and stroke symptoms that can be captured and utilized to augment the prediction of clinical stroke outcomes based on larger stroke datasets. This increase in prediction performance could then be of unique value for optimizing decisions of acute clinical care and rehabilitation approaches for individual patients.

## Data Availability

The authors agree to make the data available to any researcher for the express purposes of reproducing the here presented results and with the explicit permission for data sharing by individual sites institutional review boards.

## Acknowledgments

We are grateful to our colleagues at the J. Philip Kistler Stroke Research Center for valuable support and discussions. Furthermore, we are grateful to our research participants without whom this work would not have been possible.

## Supplemental materials

## Methods

### Neuroimaging parameters: MRI-GENIE

Neuroimages were obtained in 1T, 1.5T or 3T scanners (General Electric Medical Systems, Philips Medical Systems, Siemens, Toshiba, Marconi Medical Systems, Picker International, Inc.).

### Diffusion-weighted images (DWI)

Mostly axial orientation (2727/2770 axial, 43/2770 coronal). Axial: Reconstruction matrix 256×256mm^2^ (range: 128×128mm^2^ to 432×384mm^2^), median field-of-view 230 mm (range: 200 to 420 mm), median slice thickness 5mm (range: 2 to 7mm, gaps of 0 to 3mm), median TR 4.773ms, median TE 92ms. Coronal: reconstruction matrix 256×256 mm^2^, median field-of-view 260mm, median slice thickness 5mm, median TR 8.200ms, median TE 112ms. Mostly 3 directions (range: 3 to 25). Mostly low b-value 0s/mm^2^ (range: 0 to 50s/mm^2^), high b-value 1000s/mm^2^ (range: 800 to 2000s/mm^2^).

### Neuroimaging parameters: MGH-based cohort

Neuroimaging scans were obtained on either a Siemens (Munich, Germany) 3T MRI or a General Electric (Fairfield, CT) 1.5T MRI machine.

#### Diffusion-weighted images (DWI)

Echo time: 60 to 120 ms; repetition time: 5300–5600 ms; slice thickness: 5 mm with a 1-mm gap.

### Segmentation of stroke lesions: MGH-based cohort

The following preprocessing steps were applied: Data were resampled to 0.89 × 0.89 × 6 mm voxel spacing (the median voxel spacing of the training dataset). Data was then skull-stripped, bias corrected and normalized (image intensities with zero mean, unit variance).

We utilized a symmetrical 3D 5-level U-Net architecture with preprocessed DWI volumes as input and a probability map of the likely stroke lesions as output. Feature map downsampling and upsampling was implemented through strided convolution and trilinear interpolation, respectively. Group Normalization (with group size of 16) in lieu of Batch Normalization is used to accommodate the smaller batch size necessary to train a large patch 3D model. We used rectified linear unit (ReLU) activation in all layers, but the final one, where we used a sigmoid activation function.

We trained our network on patches of size 144×176×24 voxels with batch size 2. Patches were sampled at random during training. The network was trained to maximize the Dice Similarity Coefficient (DSC) between predicted label maps and ground truth, as expressed by the following equation:

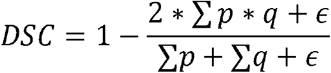

(*p* is the ground truth, *q* the predicted map, *ϵ* is used to prevent floating point instability when the magnitude of the denominator is small (set to 1)). Training relied on the SGD optimizer with decoupled weight decay, the learning rate was progressively decreased:

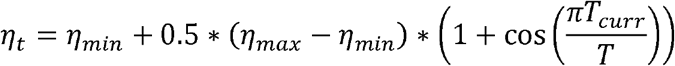

*(η*_*max*_ initial learning rate (set to 0.1), *η*_*min*_ final learning rate (set to 0.0001), *T*_*curr*_ current iteration counter, and total number of iterations to train for (set to 150 epochs)).

Furthermore, we applied a weight decay of 0.00002 to all convolutional kernel parameters, leaving biases and scales un-regularized. Real-time data augmentation was employed during the training process (random axis mirror flips (for all 3 axes), isotropic scaling (.75 to 1.25), rotations (−15° to 15°) around all three axes, and gamma corrections (.75 to 1.25), all with probability 0.5).

